# Prompt Engineering Enables Open-Source LLMs to Match Proprietary Models in Diagnostic Accuracy for Annotation of Radiology Reports

**DOI:** 10.1101/2025.10.13.25337785

**Authors:** Lau Andreas Petersen, Mathias Søndergaard Beck, Michael Brun Andersen, Jack Junchi Xu, Frederik Jager Bruun

## Abstract

**Aim:** The aim of this study was to test whether open-source Large Language Models (LLMs) can match the diagnostic accuracy of proprietary models in annotating Danish trauma radiology reports across three clinical findings.

**Materials and Methods:** This retrospective study included 2,939 radiology reports of trauma radiographs collected from three Danish emergency departments. The data were split, with 600 cases for prompt engineering and 2,339 for model evaluation. Eight LLMs, GPT-4o and GPT-4o-mini (OpenAI), and six Llama3 variants (Meta) were prompted to annotate the reports for fractures, effusions, and luxations. The reference standard was human annotations. The diagnostic performance was assessed using accuracy, sensitivity, specificity, PPV, and NPV with 95% confidence intervals.

**Results:** Prompt engineering improved the *Match*-score for Llama3-8b from 77.8% (95% CI: 74.4% – 81.1%) to 94.3% (95% CI: 92.5% – 96.2%).

GPT-4o achieved the highest overall diagnostic accuracy at 97.9% (95% CI: 97.3% – 98.5%), followed by Llama3.1-405b (97.1% (95% CI: 96.4% – 97.8%)), GPT-4o-mini (96.9% (95% CI: 96.2% – 97.6%)), Llama3-8b (96.9% (95% CI: 95.9% – 97.3%)), and Llama3.1-70b (96.0% (95% CI: 95.2% – 96.8%)). Across the three specific findings, all models performed best for fractures, whereas effusion and luxation were more prone to errors. Of the error types, Semantic Confusion was the most frequent, with 53.2% to 59.4% of misclassifications.

**Conclusion:** Small, open-source LLMs can accurately annotate Danish trauma radiology reports when supported by effective prompt engineering, achieving accuracy levels that rival proprietary competitors. They offer a viable, privacy-conscious alternative for clinical use, even in a low-resource language setting.

## Introduction

Radiology reports are a valuable source of detailed data, but their unstructured format significantly restricts their potential for secondary use in research and quality monitoring (1). Recent studies have highlighted the potential of Large Language Models (LLMs) to offer a novel, accurate, efficient, and user-friendly method for annotating radiology reports, thereby unlocking valuable information (2–4). In this study, we investigated the efficacy of both proprietary and opensource LLMs for annotating Danish-language reports from radiographs of patients with trauma.

Large proprietary models have predominantly captured research attention; however, small and open-source models offer compelling benefits crucial for the healthcare sector, including costeffectiveness, enhanced data protection, transparency, and reduced vendor lock-in (5–7). This makes them a more appealing and sustainable option for widespread clinical implementation.

Much of the existing research on large language models (LLMs) has focused on OpenAI’s GPT family of models (2,3,8,9), which, while powerful, are proprietary and often financially and computationally demanding to use. These characteristics pose significant challenges for widespread implementation in hospital environments, where budgetary constraints and paramount patient data privacy concerns limit the use of external cloud-based solutions (6). Consequently, the emergence of smaller, open-source LLMs presents a more feasible and cautious approach for healthcare applications.

Furthermore, the current literature on LLMs is heavily concentrated on high- or medium-resource languages, which are widely spoken and well-represented in training data, leaving low-resource languages critically underexplored (2,3,8–11). This linguistic disparity is particularly relevant for small languages, such as Danish, where limited model exposure during pre-training may inherently affect diagnostic accuracy, making direct evaluation essential.

Finally, although LLMs have been evaluated across diverse specialties and imaging modalities, such as oncology or MRI reports, research specifically on their application in trauma radiology remains scarce (3,11). Trauma radiology presents unique challenges due to its acute nature and highly specialized terminology. As such, it represents an important and currently underexamined area for thoroughly evaluating the performance of LLMs in a demanding clinical context.

Against this background, we aimed to test whether careful prompt engineering can enable a small, open-source LLM to match the diagnostic accuracy of state-of-the-art proprietary LLMs in annotating Danish trauma radiology reports across three clinical findings. Second, we aimed to estimate the diagnostic accuracy of eight widely used LLMs when annotating radiology reports written in Danish.

## Methods

The data used in this study were obtained as part of a separate project approved by the Institutional Review Board (ID 22070206). All data were anonymized prior to extraction, and a waiver of informed consent was granted due to the use of non-personally identifiable information.

### Study Sample

The study sample was collected for the AI Fracture project at our institution and consisted of all musculoskeletal skeletal radiographs referred from the emergency departments of three hospital centers: over 14 consecutive days at two centers and 21 consecutive days at the third. To ensure sufficient representation, the dataset was enriched with additional patients from the third center until each anatomical region included at least 50 fracture cases. For knee examinations, enrichment continued until 50 effusion cases were identified. A target of 50 positive cases per category was pragmatically set to ensure statistical robustness for the binary classification evaluation. Imaging was conducted at seven sites. Examinations were performed between January 1^st^ and January 21^st^, 2023 (third center) or January 8^th^ and January 21^st^, 2023 (the other two centers), with the latest examination related to the enrichment conducted on April 22^nd^, 2023.

### Reference Standard

All radiology reports were retrospectively reviewed by a medical doctor with three years of experience in radiology (FJB) and annotated for the presence of fractures, effusion, and luxation (each recorded as True or False). These annotations served as the reference standard against which the model’s accuracy was evaluated. All annotations were completed prior to conducting the index test; thus, the annotator was blinded to the index test outputs during the creation of the reference standard. Similarly, the LLMs did not receive any additional information beyond anonymized radiology reports.

### Selection of Large Language Models for Index Test

The selection of the included LLMs was based on two main criteria: licensing model and performance. OpenAI’s GPT-4o and GPT-4o-mini were chosen as state-of-the-art proprietary models, whereas six models from three generations of Meta’s Llama series in various sizes were selected because of their status as leading open-source models. The selected Llama-models varied in the number of parameters, ranging from Llama3.2-3b with 3 billion parameters to Llama3.1-405b with 405 billion parameters.

OpenAI’s two models and three of the six Llama-models (Llama3-70b, Llama3.1-70b, and Llama3.1-405b) were accessed via an API (www.llama-api.com and www.groq.com), while the remaining three Llama-models (Llama3-8b, Llama3.1-8b and Llama3.2-3b) were run locally.

The selected models were prompted to assess the radiology reports for the presence of a fracture, effusion, or luxation, with each condition recorded as True or False. Their outputs were subsequently compared with the reference standard established by human annotation to evaluate the model accuracy.

### Prompt Development

A range of prompt engineering techniques was evaluated on the first 600 consecutive reports from the study sample through an iterative development process designed to optimize the model accuracy and output structure. The goal was to identify a prompt that consistently produced a valid, structured JSON output while maximizing agreement with the reference annotations.

The development process involved the sequential testing of three core techniques: (1) persona prompting (e.g., instructing the model to act as a radiologist), (2) iterative refinement prompting (e.g., prompting the model to reevaluate and correct incomplete or inaccurate outputs), and (3) few-shot prompting, where examples of correctly annotated reports were included in the prompt. Combinations of these techniques were also evaluated.

For each iteration, the prompt was modified and ran on all 600 reports reserved for prompt development using the Llama3-8B model. Model performance was assessed using a binary variable, *Match*, which was defined as complete agreement between the model output and reference standard across all three target variables. Prompts yielding inconsistent JSON structures or lower *match scores* were discarded.

### Error Analysis

To gain insight into the circumstances under which the models failed, an error analysis was conducted on the outputs of four selected models: Llama3-8B, Llama3.1-405B, GPT-4o, and GPT-4o-mini. These models were chosen based on their strong overall performance, diversity in licensing models, and parameter count. All errors were independently categorized by the primary authors (LAP and MSB), and any disagreements were resolved by consensus with a medical doctor (FJB). The resulting error categories were condensed to facilitate analysis.

### Statistical Analysis

The diagnostic performance of the models was evaluated using accuracy, sensitivity, specificity, positive predictive value (PPV), and negative predictive value (NPV). Binomial distribution was used to calculate the 95% confidence intervals (CIs). All statistical analyses were conducted using RStudio (v. 2024.09.1 + 394), while all scripts for interacting with the LLMs were written in Python (v. 3.9.7), using Ollama (v. 0.5.7) to run models locally.

## Results

### Characteristics of the Study Sample

We identified 2972 free-text reports based on radiographs from trauma patients; 33 were excluded due to missing X-ray pictures (n=9), missing reports (n=4), patient age under 2 years (n=16), and technical errors (n=4). Of the remaining 2939 reports, the first 600 were reserved for prompt development and engineering, resulting in a final study sample of 2339 reports (Table 1). The mean age at examination was 43.9 (SD: ±26.4), 1228 (53%) of the patients were female.

**Table 1:**
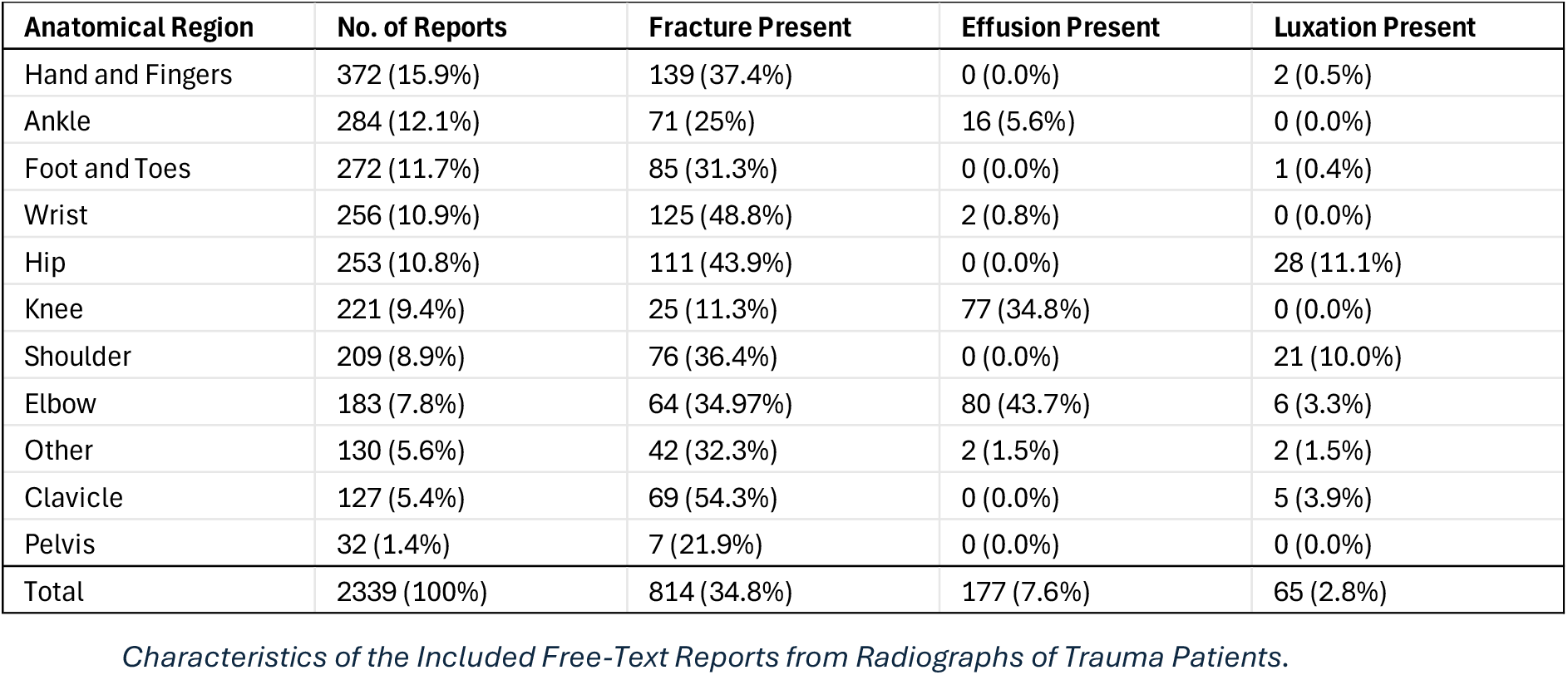
Characteristics of Free-Text Reports from Radiographs of Trauma Patients.

**Table 2:** Prompt Engineering Results. Note: The data are percentages with 95 confidence intervals in parentheses.

Results for different iterative prompt engineering strategies applied to the model. The table shows the percentage of correct Matches for each strategy iteration, with 95% confidence intervals (CI) provided in parentheses.
| Table 2: Prompt Engineering Results |  |  |  |  |  |  |  |  |
| --- | --- | --- | --- | --- | --- | --- | --- | --- |
| Note: The data are percentages with 95 confidence intervals in parentheses. |  |  |  |  |  |  |  |  |
|  | Iteration 1 | Iteration 2 | Iteration 3 | Iteration 4 | Iteration 5 | Iteration 6 | Iteration 7 | Iteration 8 |
| Prompt Strategy | Persona | Persona<br>3 few shot | Iterative<br>Refinement | Iterative<br>Refinement<br>w. Persona | Persona | Persona<br>Definitions<br>3 Few Shot | Persona<br>Definition<br>3 Few Shot | Persona<br>Definitions<br>4 Few Shots |
| Match | 77.8<br>(74.4, 81.1) | 78.8<br>(75.6, 82.1) | 69.2<br>(65.5, 72.9) | 79.2<br>(75.9, 82.4) | 85.5<br>(82.7, 88.3) | 83.7<br>(80.7, 86.6) | 90.7<br>(88.3, 93.0) | 94.3<br>(92.5, 96.2) |

Of the 18,712 outputs generated by the eight models, only one formatting error was found. This error originated from Llama3-8b, whereas the remaining models consistently produced data in the correct format.

### Prompt Engineering

Over eight iterations, three prompt engineering strategies were explored using the development subset of the study sample (n = 600). The initial ‘persona’-based prompt yielded a match score of 77.8% (95% CI: 74.4% – 81.1%). Adding few-shot examples in the second iteration improved the performance slightly to 78.8% (95% CI: 75.6% – 82.1%). An alternative approach based on iterative refinement was evaluated, producing a score of 69.2% (95% CI: 65.5 – 72.3%). A hybrid strategy combining persona prompting with iterative elements improved accuracy but introduced inefficiencies and formatting inconsistencies. Error analysis from prior runs informed a return to the persona-based approach, leading to a refined prompt that significantly increased performance to 85.5% (95% CI: 82.7% – 88.3%). Subsequent iterations incorporated few-shot examples targeted at common error types, ultimately achieving a final match score of 94.3% (95% CI: 92.5% – 96.2%).

### Overall and Finding-specific Accuracy

Figure 1 provides a visual representation of the overall diagnostic accuracy of the models, while the data for the overall accuracy and the models’ accuracy on the three specific findings (fracture, effusion, and luxation) are presented in Table 3.

**Table 3:**
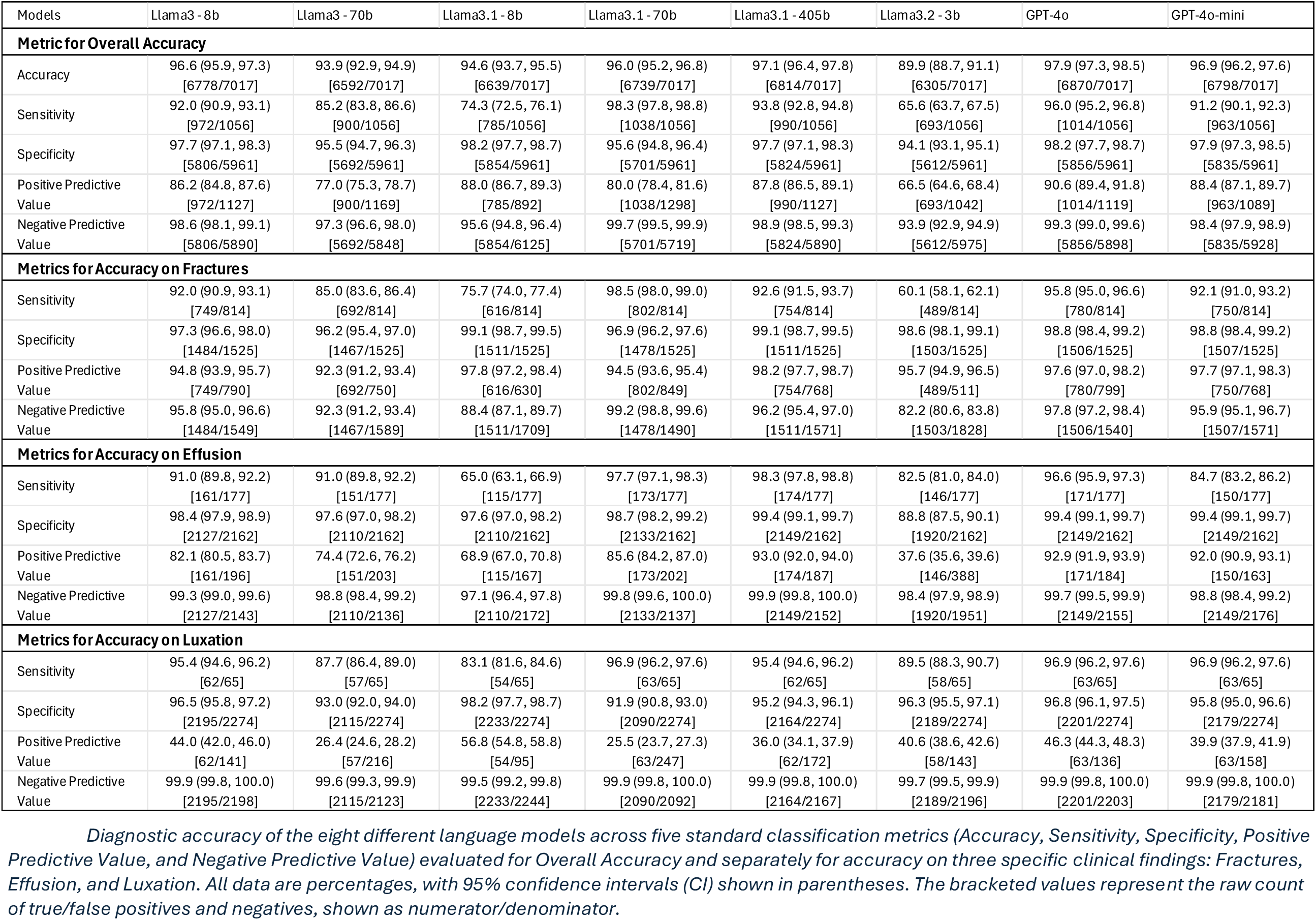
Accuracy Results. Note: The data are percentages with 95% confidence intervals in parentheses and numerators/denominators in brackets.

**Figure 1.**
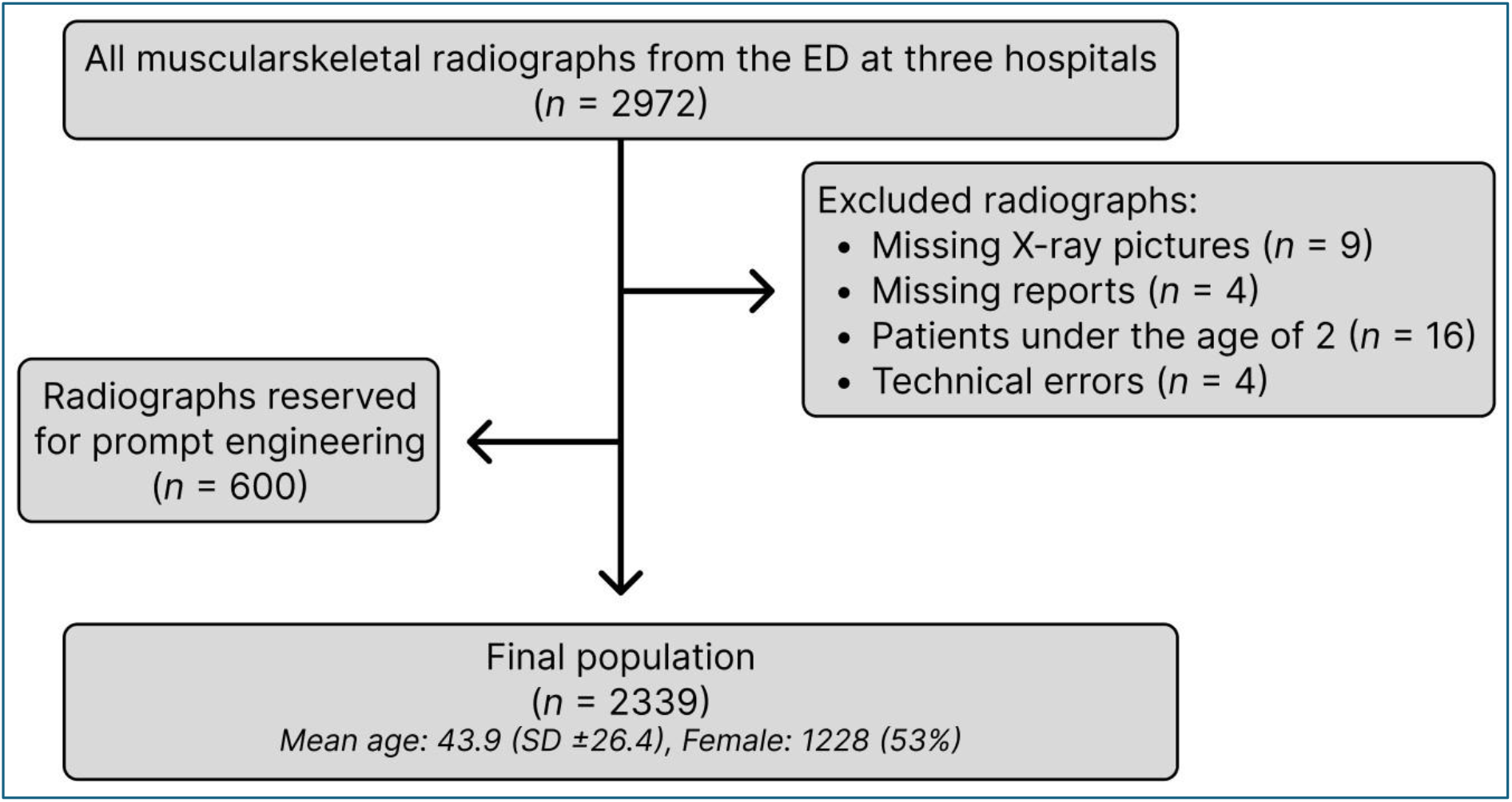
Flow Chart for Study Data

#### Overall Diagnostic Accuracy

The models achieved accuracy scores ranging from 89.9% (95% CI: 88.7% – 91.1%) for Llama3.2-3b to 97.9% (95% CI: 97.3% – 98.5%) for GPT-4o (Fig. 2, Table 3). Scores above 90% were also observed across all models for specificity and NPV, ranging from 94.1% (95% CI: 93.1% – 95.1%) to 98.2% (95% CI: 97.7% – 98.7%) and 93.9% (95% CI: 92.9% – 94.9%) to 99.7% (95% CI: 99.5% – 99.9%), respectively. However, the models generally scored lower on sensitivity, with Llama3.2-3b and Llama3.1-8b dropping to 65.6% (95% CI: 63.7% – 67.5%) and 74.3% (95% CI: 72.5% – 76.1%), respectively. More than half of the models achieved scores above 91.2% – including Llama3-8b, with larger models such as Llama3.1-70b and GPT-4o reaching scores above 95%. For PPV, GPT-4o was the only model to score above 90% – achieving 90.6% (95% CI: 89.4% – 91.8%). Across the five metrics, Llama3-8b, along with the newer and larger Llama3.1-70b, Llama3.1-405b, GPT-4o, and GPT-4o-mini, maintained scores consistently above 90%.

**Figure 2.**
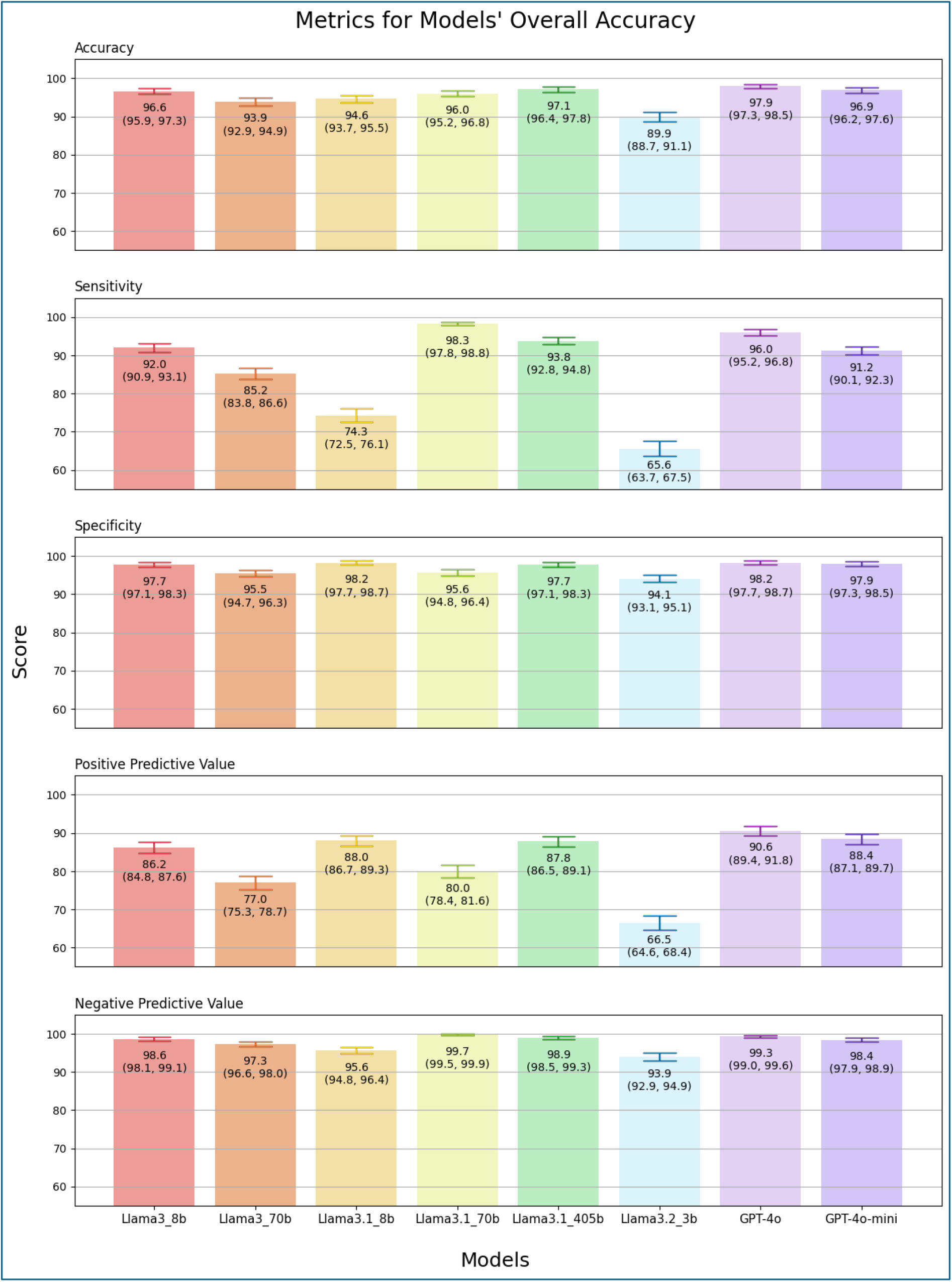
Models’ Accuracy, Sensitivity, Specificity, Positive Predictive Value (PPV), and Negative Predictive Value (NPV). Bars represent the score, with the central number indicating the score achieved and the horizontal error lines and parenthetical values indicating the 95% Confidence Interval (CI).

#### Finding-Specific Accuracy

For the correct annotation of fractures in reports, Llama3-8b, Llama3.1-70b, Llama3.1-405b, GPT-4o, and GPT-4o-mini achieved scores consistently exceeding 92% across all metrics (Fig. 3, Table 3). Llama3-8b demonstrated strong accuracy, achieving 92.0% (95% CI: 90.9%–93.1%) in sensitivity, 97.3% (95% CI: 96.6%–98.0%) in specificity, 94.8% (95% CI: 93.9%–95.7%) in PPV, and 95.8% (95% CI: 95.0%–96.6%) in NPV. In contrast, Llama3.2-3b exhibited lower accuracy than the other models, with particularly low sensitivity at 60.1% (95% CI: 58.1% – 62.1%) and a PPV of 82.2% (95% CI: 80.6% – 83.8%).

**Figure 3.**
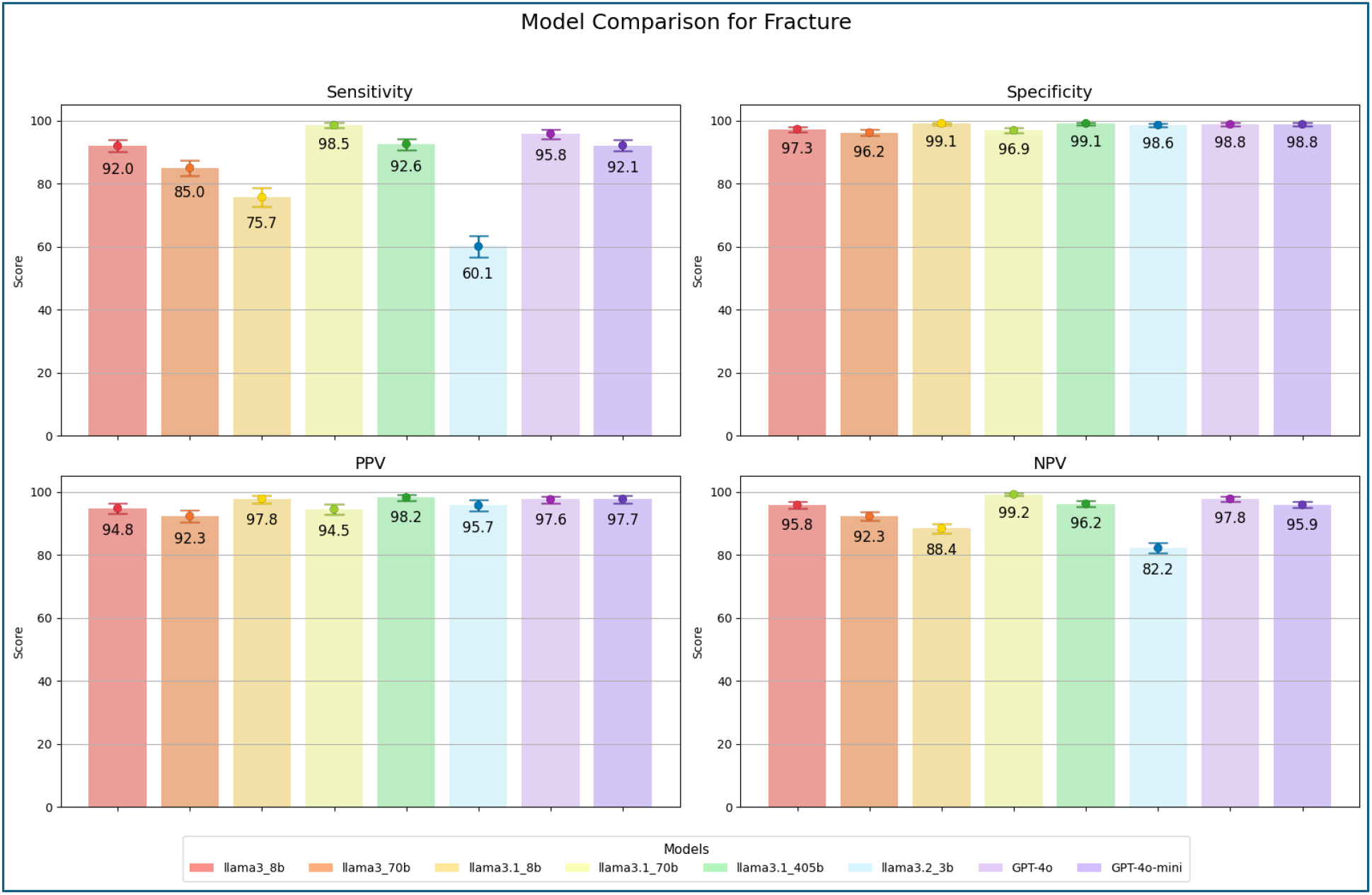
Model performance comparison for Fracture classification, showing Sensitivity, Specificity, PPV, and NPV scores with 95% CIs.

For effusions, Llama3.1-70b, Llama3.1-405b, GPT-4o, and GPT-4o-mini achieved overall accuracy exceeding 90% (Fig. 4, Table 3). Llama3-8b followed closely, with a lower PPV of 82.1% (95% CI: 80.5%–83.7%). Llama3.1-8b and Llama3.2-3b achieved the lowest accuracy scores for effusion detection, with PPV scores of 68.9 % (95% CI: 67.0 % – 70.8 %) and 37.6 % (35.6 % – 39.6 %), respectively.

**Figure 4.**
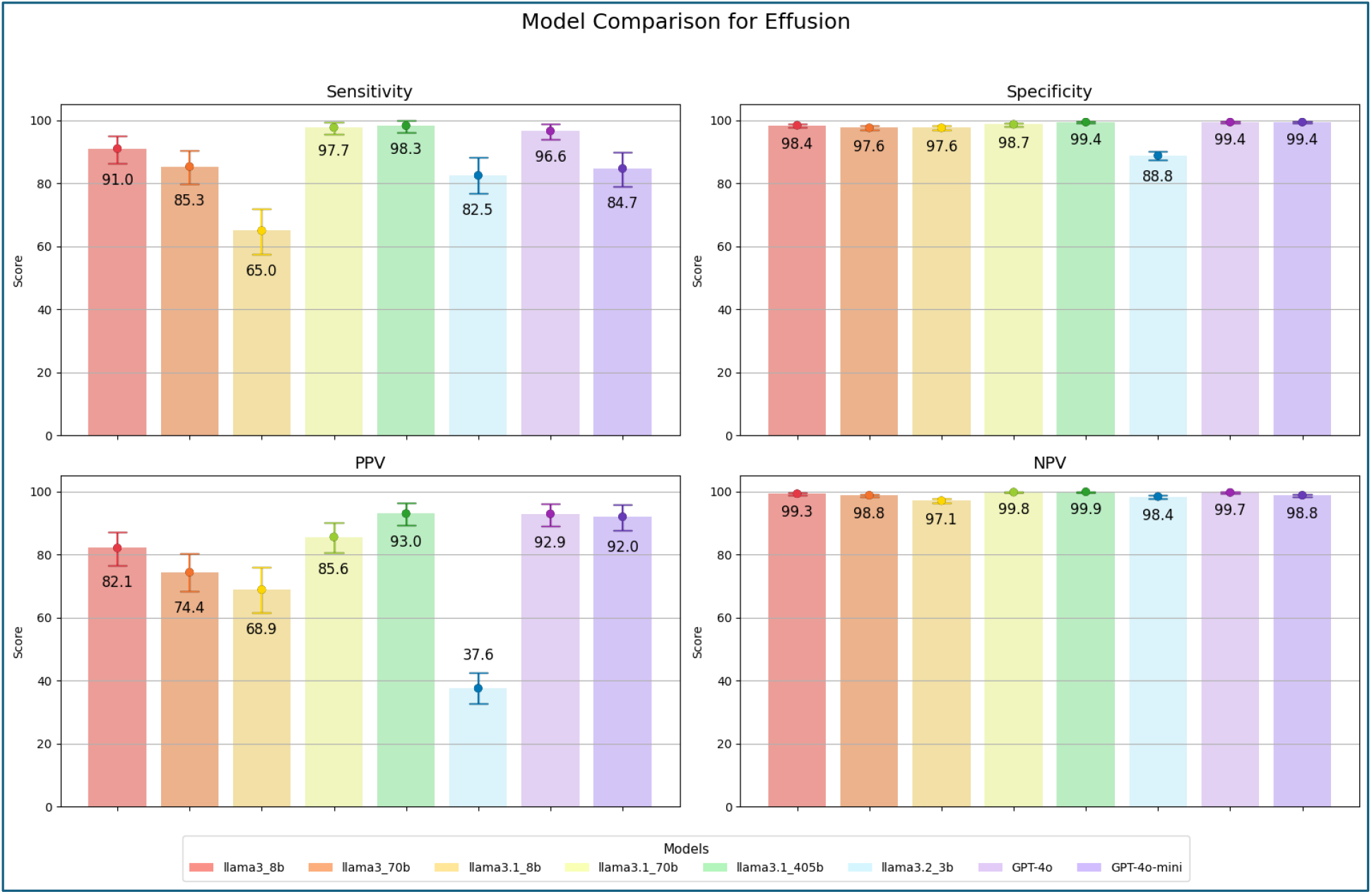
Model performance comparison for Effusion classification, showing Sensitivity, Specificity, PPV, and NPV scores with 95% CIs.

As with the annotation of effusion and fracture, Llama3-8b, Llama3.1-70b, Llama3.1-405b, GPT-4o, and GPT-4o-mini achieved high sensitivity, specificity, and NPV scores, all exceeding 95% (Fig. 5, Table 3). However, all models exhibited low PPV for luxation, with Llama3.1-8b being the only model to exceed 50% – achieving a PPV of 56.8% (95% CI: 54.8%–58.8%).

**Figure 5.**
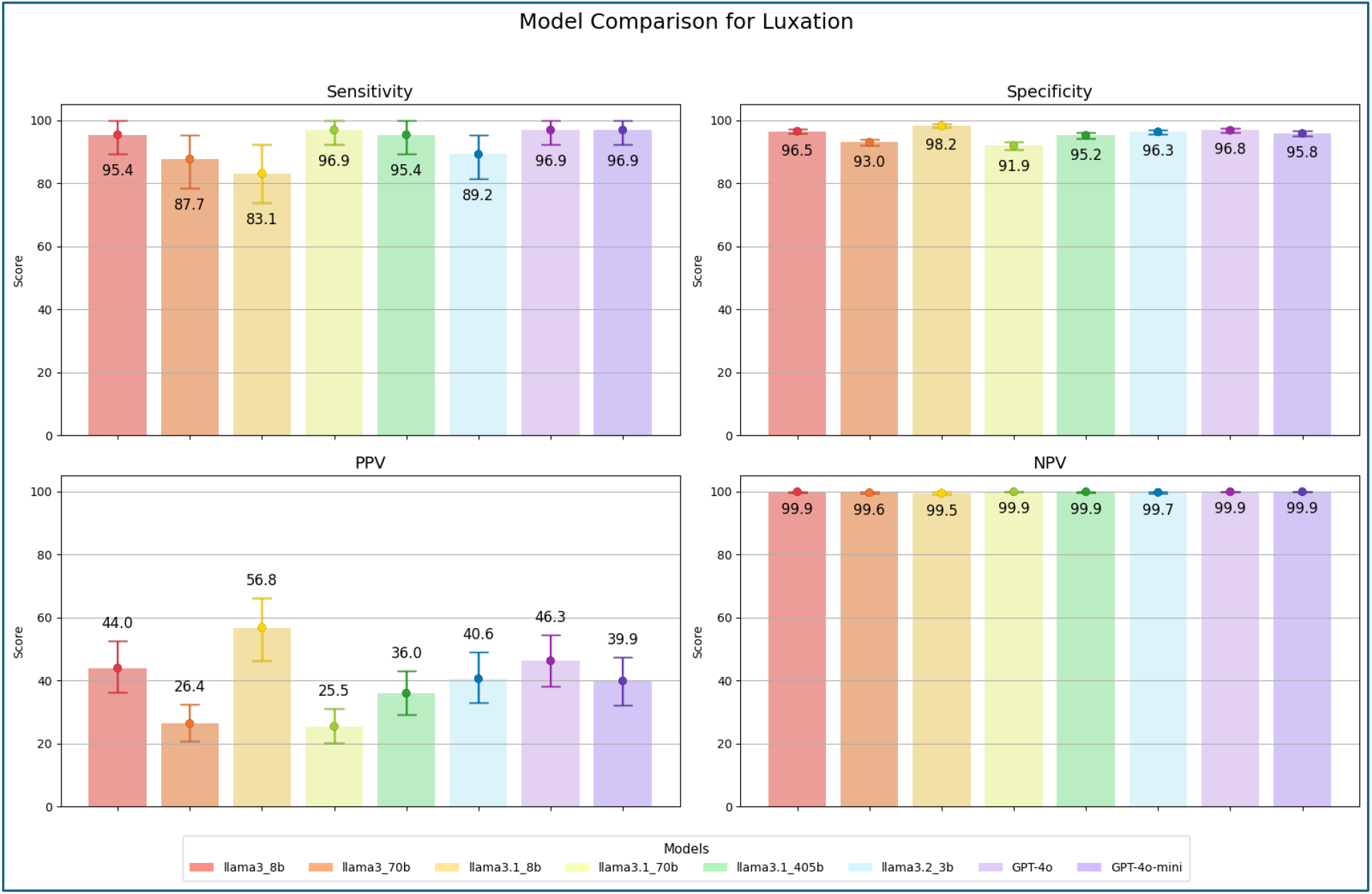
Model performance comparison for Luxation classification, showing Sensitivity, Specificity, PPV, and NPV scores with 95% CIs.

### Error Types

Semantic Confusion accounted for the majority of errors across the four selected models, with GPT-4o-mini having the largest fraction of Semantic Confusion errors of all the four models at 59% (Fig. 6). The following two most common error types were Report Ambiguity and Unrecognized Synonyms, ranging from 10.9% to 14.4% and 12.6% to 18.3% – respectively. The fourth most frequently occurring error type was Incorrect Annotation, while the fifth most frequently occurring error type was Healed Findings. Finally, a group of errors was classified as ‘Unclassified Errors’, accounting for 14 of the total 631 errors.

**Figure 6.**
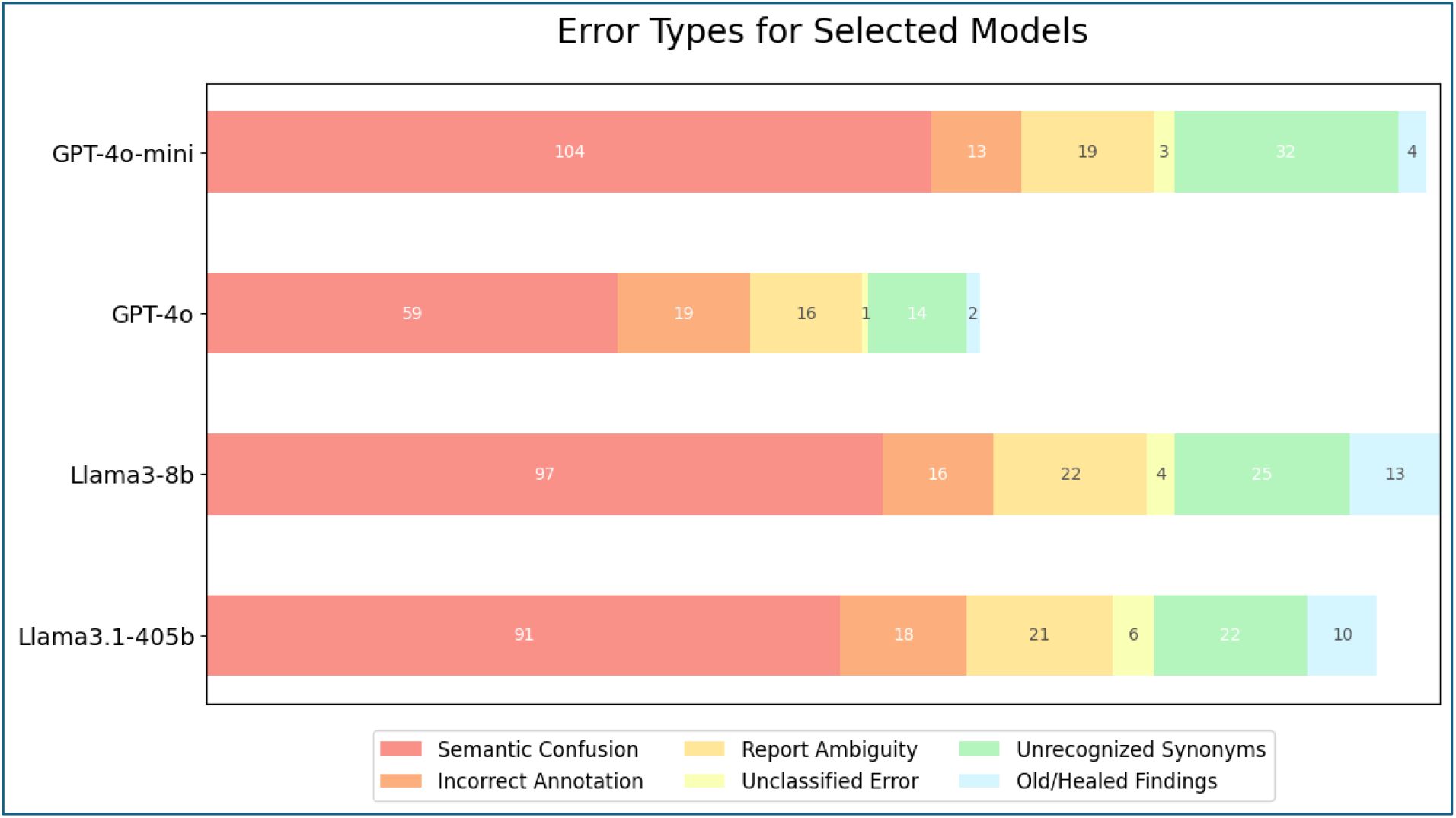
Distribution of Error Types for Selected Models.

## Discussion

This study aimed to assess whether LLMs can effectively structure such data by annotating realworld trauma reports from Denmark. Our results demonstrate that LLMs, including small opensource models, can achieve high diagnostic accuracy.

Remarkably, the smaller Llama models, specifically Llama3-8b and Llama3.1-70b, performed on par with their larger and/or proprietary counterparts. This finding is particularly significant given their lightweight nature and suitability for local deployment, which directly addresses critical concerns regarding cost-effectiveness, data privacy, and vendor lock-in in healthcare applications.

Our findings align with and contribute to the understanding of the applications of LLMs in radiology. The general utility of LLMs in extracting structured information from clinical text is well-established (12–14), and our study further substantiates this potential within the specific context of trauma radiology and low-resource languages.

A key contribution of this study is the direct comparison of proprietary and open-source LLMs. While much early research focused on large proprietary models such as OpenAI’s GPT series, our results show that smaller open-source Llama models (Llama3-8b and Llama3.1-70b) performed as well as their larger proprietary counterparts. This is crucial for healthcare adoption, aligning with emerging evidence that open-source alternatives offer cost-effectiveness, data privacy, and reduced vendor lock-in, without compromising diagnostic accuracy (15,16). These findings are consistent with recent studies, such as Li et al., who similarly observed that proprietary and open-source models perform comparably, despite generally lower overall accuracy than reported in our results (17). A similar pattern was noted by Dorfner et al. in their analysis of chest radiograph reports: while the proprietary model (GPT-4) initially outperformed the opensource models off-the-shelf, the performance gap narrowed substantially with the application of effective prompt engineering (18).

Furthermore, our focus on a low-resource language addresses a significant gap. Our successful implementation, which achieved high accuracy through careful prompt engineering, demonstrates that effective adaptation strategies can overcome these language barriers. To the best of our knowledge, no studies have applied LLMs in the same manner as presented here, our results are nonetheless in line with broader findings on LLM performance in low-resource language settings (19,20).

Finally, our investigation of trauma radiology reports highlights an underexplored domain. The specialized terminology and acute nature of these reports present unique challenges to translators. Our results confirm that with effective prompt design, LLMs are robust enough to accurately interpret complex, domain-specific information, thereby expanding their known utility across diverse medical specialties.

The majority of errors across the four selected models (Llama3-8B, Llama3.1-405B, GPT-4o, and GPT-4o-mini) stemmed from Semantic Confusion, comprising between 53.2% and 59.4%. %. An example of semantic confusion is when the word “dislocated” is used to describe a fracture in which two bone fragments are misaligned. Models incorrectly interpret the presence of “dislocated” as a luxation, which is the technical term for a bone misaligned from its position inside a joint. This suggests that LLMs struggle with the nuanced phrasing and contextual subtleties common in clinical prose.

Report Ambiguity (10.9% to 14.4%) and Unrecognized Synonyms (12.6% to 18.3%) were the second and third most frequent error types, respectively, highlighting challenges associated with vague descriptions and varied medical terminology. These issues are consistent with the observations of Meddeb et al. (19), who also identified limitations in large language models when processing specialized medical vocabulary. Errors due to Incorrect Annotation and Healed Findings were also present, with the latter particularly revealing the models’ difficulty in distinguishing temporal aspects, such as “fresh” versus “old” fractures.

These clearly categorized error types provide actionable insights for further refinement. Future research should explore advanced prompt engineering techniques that specifically target semantic nuances and temporal cues. Additionally, domain-specific fine-tuning or employing retrieval-augmented generation (RAG) could significantly mitigate the identified error patterns. Understanding these specific failure modes transforms them into valuable signposts for developing more robust and clinically reliable LLMs.

While the findings offer valuable insights, they must be viewed in light of certain limitations that affect their scope and reliability. This study relied on a prompt developed specifically for the Llama3-8b model, which might limit its generalizability to other model architectures. It is possible that a prompt designed for a different model, such as GPT-4o-mini, might perform differently or even enhance accuracy within certain model families. Interestingly, the GPT-4 models might be more versatile, as they performed on par with or better than the Llama models, despite the prompt being developed on Llama3-8b. This suggests that GPT-4 models may be more adaptable to prompts that are not explicitly optimized for them. Additionally, as new models and architectures continue to emerge, the findings of this study may not fully extend to future systems, further limiting their general applicability.

Moreover, the evaluation was confined to three specific findings (fracture, effusion, luxation) and musculoskeletal radiographs from emergency departments, which may limit the generalizability to other conditions, imaging modalities (e.g., MRI, CT), or clinical settings. While a strength for “low-resource” languages, the single-language focus of Danish also means that our findings may not directly translate to other languages without further validation. Finally, although a medical doctor established the reference standard, the use of a single annotator could introduce subtle biases, and inter-rater reliability was not assessed.

In conclusion, our study demonstrates that LLMs, particularly well-engineered open-source models, are feasible for high-accuracy annotation of Danish trauma radiology reports. This promises significant efficiency gains for research and quality control by structuring valuable, otherwise inaccessible free-text data. Future work should expand to other languages and modalities, integrate these tools into clinical or research workflows, and refine models to address identified error types to accelerate LLM adoption in real-world healthcare settings.

## Data Availability

Data are available upon reasonable request to the corresponding author

